# Racial differences in lifetime healthcare costs associated with obesity-related multimorbidity among the U.S. population aged 40 years or older

**DOI:** 10.64898/2026.08.09.26360041

**Authors:** Preeti Pushpalata Zanwar, Mei Wang, Nathan Logan, Su-Hsin Chang

**Author notes:** **Corresponding author:** Su-Hsin Chang, SM PhD, Division of Public Health Sciences, Department of Surgery Washington University School of Medicine, 660 S. Euclid Avenue, Campus Box 8100 St. Louis, MO 63110.

## Abstract

**Background:** Obesity is a costly cause of morbidity and disability. Previous research has well documented that Black persons bear a disproportionate financial burden of obesity-related multimorbidity (ORM); however, healthcare costs have not been examined over the lifespan.

**Objective:** Among U.S. general population age ≥40 years, we quantified racial difference in 1) lifetime healthcare costs (LHC’s), and 2) lifetime healthcare cost differential (LCD) associated with ORM.

**Methods:** We used data from the Medical Expenditure Panel Survey, 2008-2012, and targeted four obesity-related diseases (ORDs): diabetes, hypertension, coronary heart disease, and stroke. A published Markov model was used to model individuals’ life histories of ORDs and to compute LHC’s. LCD associated with ORM was computed by taking the difference between LHC for those with ORM and LHC for members without ORD. Racial differences were quantified by taking the differences in LHC or LCD between White and Black men/women.

**Results:** We included 53,035 Black and White individuals representing 97,229,611 (se=2,104,365) U.S. Black (12.4%) and White (87.6%) populations aged ≥40 years. ORM was more prevalent in the Black (21.2%) than the White populations (13.4%). Racial difference in LHC’s for women/men with ORM and LCDs associated with ORM (2012$) was $31,035/43,595 and $11,350/26,948 for age 40-49, $21,567/25,615 and $3,846/9,808 for 50-59, $9,863/18,515 and −$2,566/7,426 for 60-69, −$8,220/16,285 and −$11,524/3,865 for 70-79.

**Conclusions:** Racial differences in LHC’s and LCD associated with ORM persist and the scale varies by subpopulation. Future interventions designed to prevent/manage ORM are crucial to prioritize populations burdening with high LHC’s and health disparities.

**Highlights:** Racial disparities in the prevalence of obesity and obesity-related multimorbidity (ORM), and financial burden of managing ORM are well-established. We quantified racial differences in lifetime healthcare costs associated with ORM to inform future interventions and policies aiming at reducing health disparities.

## INTRODUCTION

Obesity is the second-leading cause of preventable death in the United States and an important risk factor for the development of multimorbidity (co-occurrence of ≥2 chronic conditions) and disability around the world.^1–6^ In the United States, increasing life expectancy and an aging population have contributed to increasing rates of multimorbidity.^2,4^ Although age has been a well-documented driver of multimorbidity, past studies have also investigated the relationship between gender, race, and the development of multimorbidity and found that women, Black and non-Hispanic White populations have the highest prevalence of multimorbidity in the United States among racial/ethnic groups.^6,7^ However, multimorbidity likely occurs at an earlier age in Black populations compared with Hispanic or non-Hispanic White populations.^6,8^

Prior studies, including studies by our team, have documented disparities in the prevalence of obesity, obesity-related multimorbidity (ORM), and life-years lost associated with ORM between White and Black populations in the United States.^8,9^ Black Americans, the largest racial minority group in the United States, bear a disproportionate burden of obesity and ORM.^10–12^ Increased prevalence of hypertension and diabetes in the Black population confers a higher risk of stroke and stroke mortality compared to their White counterparts.^13–15^ Studies also found that Black persons had higher incidence of fatal coronary heart disease compared to White persons.^16,17^ Additionally, the prevalence of cardiovascular- and diabetes-related hospitalizations is higher among Black populations compared to other racial groups.^18^

Financial burden of overweight, obesity, and its comorbidities are substantial and vary according to race, gender and age.^19^ In a study of MEPS data from 2008 to 2012, annual healthcare expenditures associated with higher body mass index (BMI) were three times more among those with diabetes than those without diabetes among U.S. adults ≥50 years.^20^ As black populations were disproportionately affected by obesity and ORM, we hypothesized that the financial burden of ORM in Black populations would be higher than their White counterparts. However, such difference in the financial burden of ORM may not be reflected in lifetime healthcare costs (LHC’s) as the LHC’s are determined by disease burden, access to care, and life expectancy.

While prior research has examined the healthcare costs of obesity, few studies have considered lifetime costs of ORM and racial difference in LHCs associated with ORM. The objective of our study was to quantify racial difference in 1) LHC by age, gender, and ORM status, and 2) LHC differential (LCD) associated with ORM by age and gender among U.S. general population age 40 years or older. Age ≥40 was used because ORM is prevalent in middle-aged adults.

## METHODS

### Data

We used data from the Household Component of the Full Year Consolidated Data of the Medical Expenditure Panel Survey (MEPS-HC), 2008-2012.^21^ The MEPS-HC was used to form our study population and estimate annual healthcare costs. MEPS is a set of large-scale surveys that provides nationally representative estimates of healthcare use, expenditures, sources of payment, and insurance coverage for the U.S. civilian non-institutionalized community-living population.^21^ MEPS provides the most complete data on these economic indicators compared to other surveys.^22^ MEPS-HC questionnaires are given to individual household members and their medical providers.^21^

### Study sample and population

The study population has been described previously.^23^ Our overall approach involved pooling 5-years of MEPS-HC data as recommended by MEPS analytic guidelines.^24^ This gave us a sample size (n) of 168,214 respondents after excluding non-respondents. For years 2008-2012, the MEPS non-response rate was 59.3% in 2008, 57.2% in 2009, 53.5% in 2010, 54.9% in 2011, and 56.3% in 2012. Survey non-responses over time were adjusted for in the weight variable.^25^

We included individuals who were 40-79 years in MEPS-HC, 2008-2012 (n=65,162). Individuals aged ≥80 years were not included because the MEPS top coded individuals aged ≥85 years. We further restricted our sample to non-pregnant persons (n=64,994), those without any cancer diagnosis (n=57,471),^26,27^ and those with BMI ≥18.5 (n=55,211). We further excluded individuals with age of target disease diagnosis older than their age at survey, and those with age of diagnosis <18 years. For individuals self-reporting a diagnosis of diabetes, those who reported using insulin before age 30 were also excluded to exclude those with type-1 diabetes.^23,28^ Our study sample comprised 53,035 individuals representing an average population size (N) of 97,229,611 of non-institutionalized middle-aged and older U.S. community adults for the pooled period from 2008-2012 (see Supplementary Figure 1).

### Race

Race variables available in the MEPS-HC, RACEX in years 2008-2011 and RACEV1X in year 2012 was retrieved and classified into two categories: White and Black. Ethnicity was not taken into account in this study due to insufficient data for the full range of studied disease combinations for some racial/ethnic minorities.^23^

### Studied obesity-related diseases

The target ORDs were diabetes, hypertension, coronary heart disease (CHD), and stroke, all of which are the most common chronic conditions for Medicare beneficiaries.^29^ These conditions are also among the 20 chronic conditions defined by the U.S. Department of Health and Human Services Interagency Workgroup on Multiple Chronic Conditions, and their data are available in the MEPS.^6^ In MEPS-HC, respondents who answered “*Yes*” to the following question were classified as having the disease: “*Have you ever been told by a doctor or health professional that you have diabetes/hypertension/CHD/stroke?*” Individuals who reported having a disease were then asked their age at first diagnosis.

### Other variables

Age was constructed using AGE[year]X variable from MEPS-HC, which was computed from date of birth and denotes exact age for an individual at the end of a corresponding year. Pregnancy was discerned from PREGNTX variable, which asked respondents “*If any female persons in the family had been pregnant anytime in any of the rounds of the interview?*” In our study, this question only applied to women ages ≤ 44 years as pregnant women older than 44 years were coded inapplicable in MEPS. Cancer diagnosis was ascertained from CANCERDX which asked adult persons “*whether the person had ever been diagnosed as having cancer or a malignancy of any kind”*. BMI for men and women ≥40 years was divided into three mutually exclusive categories (normal weight: BMI 18.5-24.9 kg/m^2^, overweight: BMI 25.0-29.9 kg/m^2^, and obese: BMI ≥30.0 kg/m^2^) based on BMINDX53 variable, which was calculated from self-reported height and weight.^34^ No socioeconomic variables, such as education or income, were used because we sought to study the combined cost difference associated with race as collected and reported in the MEPS survey data by age and gender.

### Annual healthcare cost

Annual healthcare costs were total expenditures used for healthcare services during that year, defined as the sum of direct payments for care provided. These include out-of-pocket payments and payments by private insurance, Medicaid, Medicare, and other sources. Payments for over-the-counter drugs are not collected in MEPS. Indirect payments not related to specific medical events, such as Medicaid Disproportionate Share and Medicare Direct Medical Education subsidies, are also not included.^22^ All dollar values were presented in 2012 U.S. dollars based on the Personal Health Care Expenditure Price Index.

Because 11.3% of the cohort had zero healthcare costs, we used a two-part model composed of a logit model in the first part estimating the probability of incurring any costs and a generalized linear model with log-link and gamma-variance function in the second part evaluating the mean costs conditional on having incurred any as described and applied previously.^30–33^ In this two-part model, we controlled for race (White, Black, and Other), age (40-49, 50-59, 60-69, 70-79 years), BMI group (normal weight, overweight, obese), any pre-existing diabetes, hypertension, CHD, or stroke, duration from time at diagnosis to time at survey, and their duration squared.

### The Markov model

To model an individual’s life history of disease progression and ORM development and to predict LHC, we used an established and calibrated Markov model with 17 mutually exclusive health states (no ORD, any combination of the four targeted ORDs, and death). This model has been described previously.^10^

For each individual in the subpopulation, the LHC was computed by the present value of the accumulated annual healthcare cost based on the age, race, gender, and health state of the individual each year. Future cost was discounted at an annual rate of 3%.^34^ The LHC for the subpopulation was computed by the mean of the LHC’s for all of the individuals in the subpopulation. For each age-race-gender-ORD status subpopulation, bootstrapping was performed 500 times to obtain means and standard errors of LHCs.

### Racial differences in LHCs by age, gender, and ORM status

We computed LHCs for populations with no ORD, one of the four targeted ORDs, and ORM. We further computed the racial difference in LHCs by taking the difference in LHCs associated with ORM between the White and the Black populations: LHC_W_-LHC_B_ by age, gender, and ORM status: no obesity-related disease (no ORD), only one obesity-related disease (1 ORD), and ORM (≥2 ORDs).

### Racial differences in lifetime cost differential (LCD) associated with ORM by age and gender

LCD associated with ORM for a subpopulation was computed by taking the difference between LHC for members with ORM and LHC for members with no ORD. We also computed the difference in LCD between White and Black populations: LCD_W_-LCD_B_ by age and gender.

### Statistical analyses and simulations

Following the analytic guidelines provided with the datasets we pooled multiple years of the data and adjusted for the complex sampling designs in all estimations and simulations.^24,35–37^ Statistical analyses for estimating annual healthcare cost as well as simulation were performed on the entire study population. However, we only present the results for the Black and the White populations to compare racial differences.

For the simulation, we present the results by age (40-49, 50-59, 60-69, 70-79), gender (women, men), race (White, Black), and ORM status (no ORD, 1 ORD, and ORM). To test differences between two races, we utilized the following statistical tests: (1) Chi-square tests for categorical variables and t-tests for continuous variables in unadjusted analyses, and (2) two-sampled t-tests with unequal variance for LHC’s and LCDs. All tests were two-sided. Statistical significance was determined using α=0.05.

Stata/SE 16 (StataCorp, College Station, TX) was used to obtain estimates of lifetime healthcare costs and costs differential for disease. MATLAB R2020a (MathWorks, Natick, MA) was used to perform simulations and bootstraps.

## RESULTS

Our sample was comprised of 53,035 Black and White individuals representing an estimated population of 97,229,611 (se=2,104,365) U.S. Black and White adults aged 40 years or older, 2008-2012 (Table 1). Among this population, 87.6% were White, and 49.5% were men. The Black and White populations were different in gender (p=0.0047), age (p<0.0001), BMI category, ORM status, and annual healthcare costs (p<0.0001). Among the White population, men and women were equal in proportion (50.0% each), while the Black population had higher proportion in women (54.7%) than men (45.3%). Overall, the proportion of the population decreased with age, but the decreasing trend was more prominent in the Black population than in the White population (40-49 years: 37.8 vs 34.3%, 50-59 years: 34.8 vs 34.0%, 60-69 years: 19.5 vs 21.4%, 70-79 years: 7.9 vs 10.3%). Compared to the White population, higher proportion of the Black population were obese (White: 33.9% Black: 44.6%). A higher proportion of the White population in comparison to the Black populations had no ORD (55.5 vs. 40.1%) whereas a higher proportion of Black populations in comparison to White populations had 1 ORD (38.7 vs. 31.1%) or had ORM (21.2 vs. 13.4%). The mean annual healthcare cost was $5,142 (SE: 87) for the White population compared to $5,276 (SE: 186) for the Black population.

**Table 1:** Characteristics of the U.S. population stratified by race, MEPS, 2008-2012, Sample size (n) = 47,557, total population (N) =97,229,611.

|  | **Total** | **White**<br>**Population** | **Black**<br>**Population** | **p-value** |
| --- | --- | --- | --- | --- |
| Sample Size, n | 47,557 | 36,917 | 10,640 |  |
| Population Size, N | 97,229,611 | 85,155,922 | 12,073,688 |  |
| Population, % | 100 | 87.6 | 12.4 |  |
| Gender, % |  |  |  | 0.0047 |
| Men | 49.5 | 50.0 | 45.3 |  |
| Women | 50.5 | 50.0 | 54.7 |  |
| Age, % |  |  |  | <0.0001 |
| 40–49 years | 34.7 | 34.3 | 37.8 |  |
| 50–59 years | 34.1 | 34.0 | 34.8 |  |
| 60–69 years | 21.2 | 21.4 | 19.5 |  |
| 70–79 years | 10.0 | 10.3 | 7.9 |  |
| BMI category, % |  |  |  | <0.0001 |
| Normal weight | 27.0 | 28.1 | 19.4 |  |
| Overweight | 37.7 | 38.0 | 36.0 |  |
| Obese | 35.2 | 33.9 | 44.6 |  |
| ORM status, % |  |  |  |  |
| No ORD | 53.6 | 55.5 | 40.1 | <0.0001 |
| 1 ORD | 32.0 | 31.1 | 38.7 |  |
| ORM | 14.3 | 13.4 | 21.2 |  |
| Annual healthcare costs, 2012\$ | | | | <0.0001 |
| Mean | 5,158.9 | 5,142.3 | 5,275.6 |  |
| Standard error | 80.0 | 87.1 | 185.8 |  |

BMI, body mass index; ORD, obesity-related disease, including hypertension, diabetes, coronary heart disease, or stroke; ORM, obesity-related multimorbidity.

Overall, women had higher LHCs than men within the same age group and with the same ORM status (Figure 1 Panel A); LHCs decreased with age but increased with number of ORDs; and other things being equal (same gender, age group, and ORM status), LHCs for the White populations were higher than those for the Black populations. Among all gender-age-ORD status-race groups, LHCs were highest in White women ages 40-49 years with ORM ($267,899); on the contrary, LHCs were lowest in Black men ages 70-79 years without ORD ($38,771).

**Figure 1.**
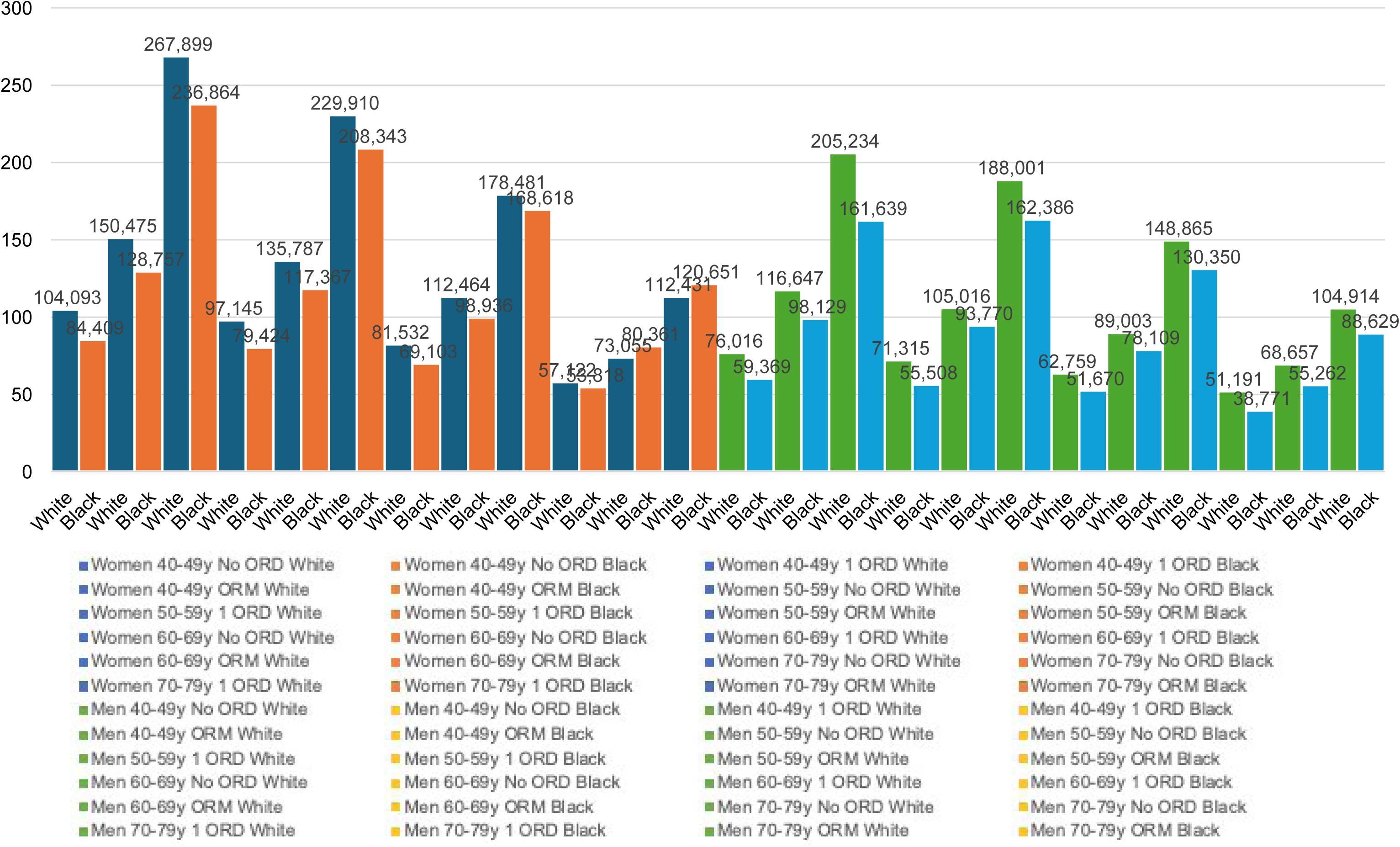
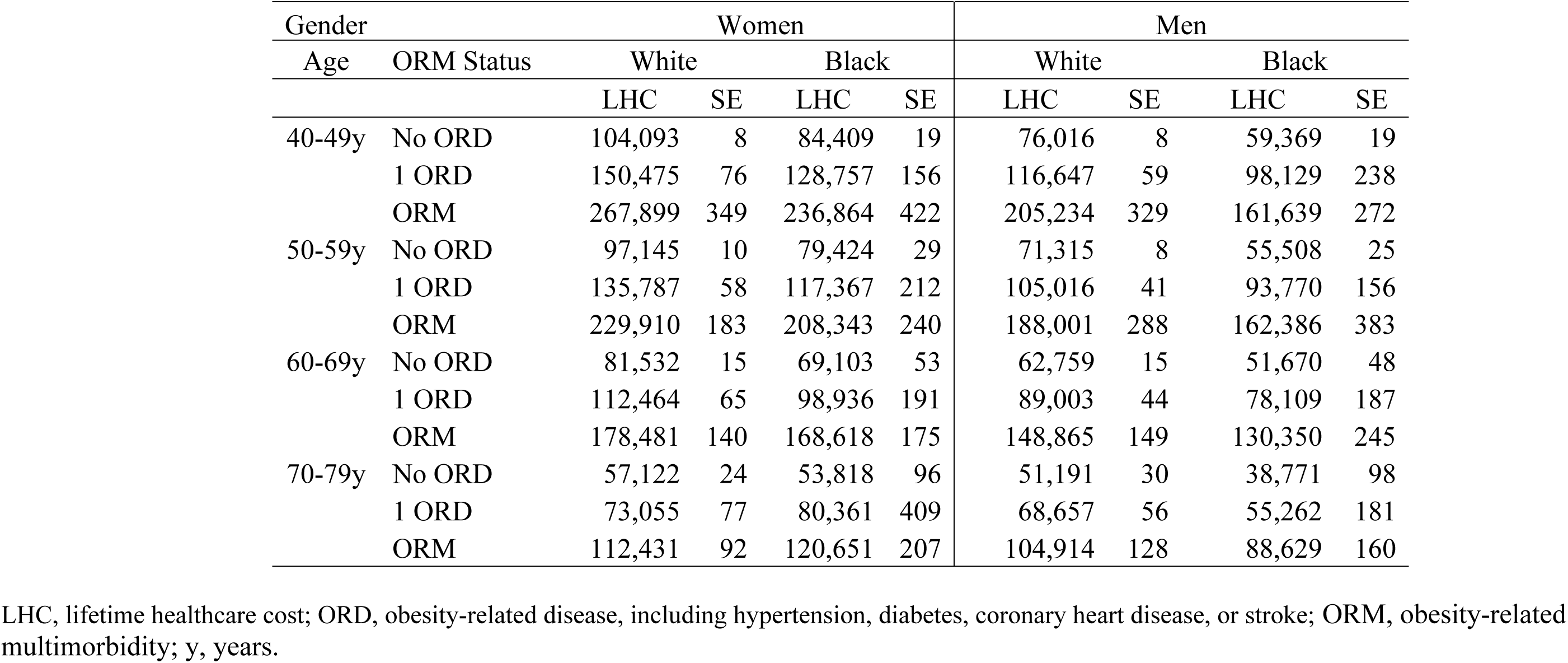
Panel A. LHC’s by age, gender, ORM status, and race.

**Figure 1.**
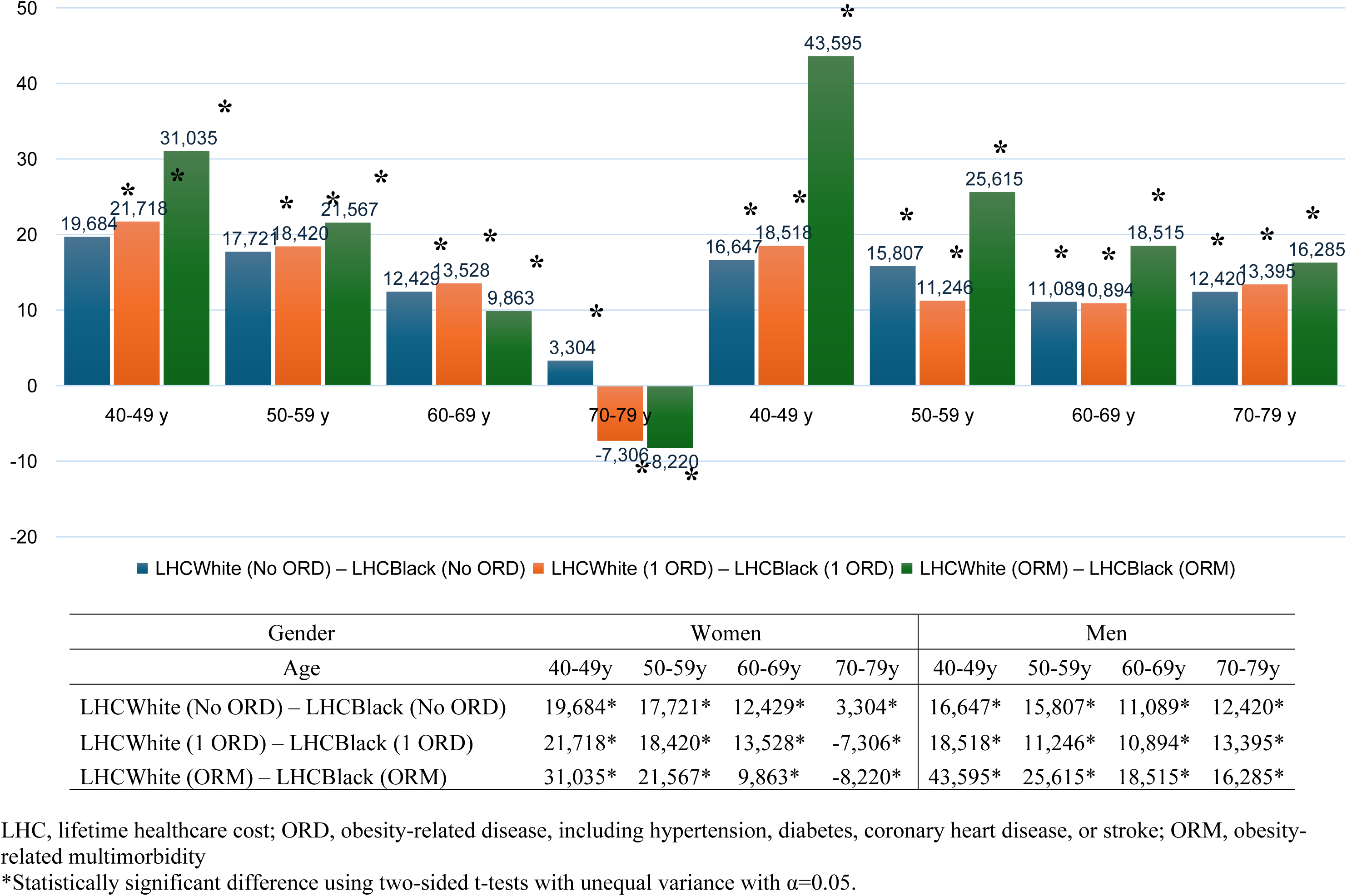
Panel B. Racial differences in LHCs by age, gender, and ORM status.

Across ORM status, racial difference in LHCs was highest in the populations with ORM for both women and men in all age groups, except for women aged 60-69 years ($9,863 for ORM <$12,429 for no ORD or $13,528 for one ORD). In fact, Black women aged 70-79 with ORM incurred higher LHC than their White counterparts, i.e., LHC_W_-LHC_B_=-$8,220, although negative, the effect size was still the largest for those with ORM among all three groups of ORD status. In the same age group, Black women with one ORD also incurred higher LHC than their White counterparts, i.e., LHC_W_-LHC_B_=-$7,306, while this does not hold for men in this age group. Racial differences in LHCs were statistically significant in all gender-age-ORD status groups. Furthermore, across all gender-age-ORD status groups, racial difference in LHCs was the highest in men aged 40-49 years with ORM ($43,595), followed by women in the same age group with ORM ($31,035) and men aged 50-59 years with ORM ($25,615).

In women, LCD associated with ORM decreased by age in both racial groups (Figure 1, panel A): White: $163,806 for ages 40-49, $132,765 for ages 50-59, $96,949 for ages 60-69, and $55,309 for ages 70-79; Black: $152,456 for ages 40-49, $128,919 for ages 50-59, $99,515 for ages 60-69, and $66,833 for ages 70-79. In men this trend only held for White populations ($129,218 for ages 40-49, $116,687 for ages 50-59, $86,106 for ages 60-69, and $53,723 for ages 70-79), but not for Black populations ($102,270 for ages 40-49, $106,878 for ages 50-59, $78,680 for ages 60-69, and $49,858 for ages 70-79).

Racial differences in LCD associated with ORM were statistically significant in all gender-age groups. In men, LCD associated with ORM was higher in White populations than Black populations across all age groups with the LCD_W_-LCD_B_ >0 (Figure 2, panel B), while in women, this is true for the younger age groups (40-49 and 50-59 years), not the older age groups (60-69 and 70-79 years).

**Figure 2.**
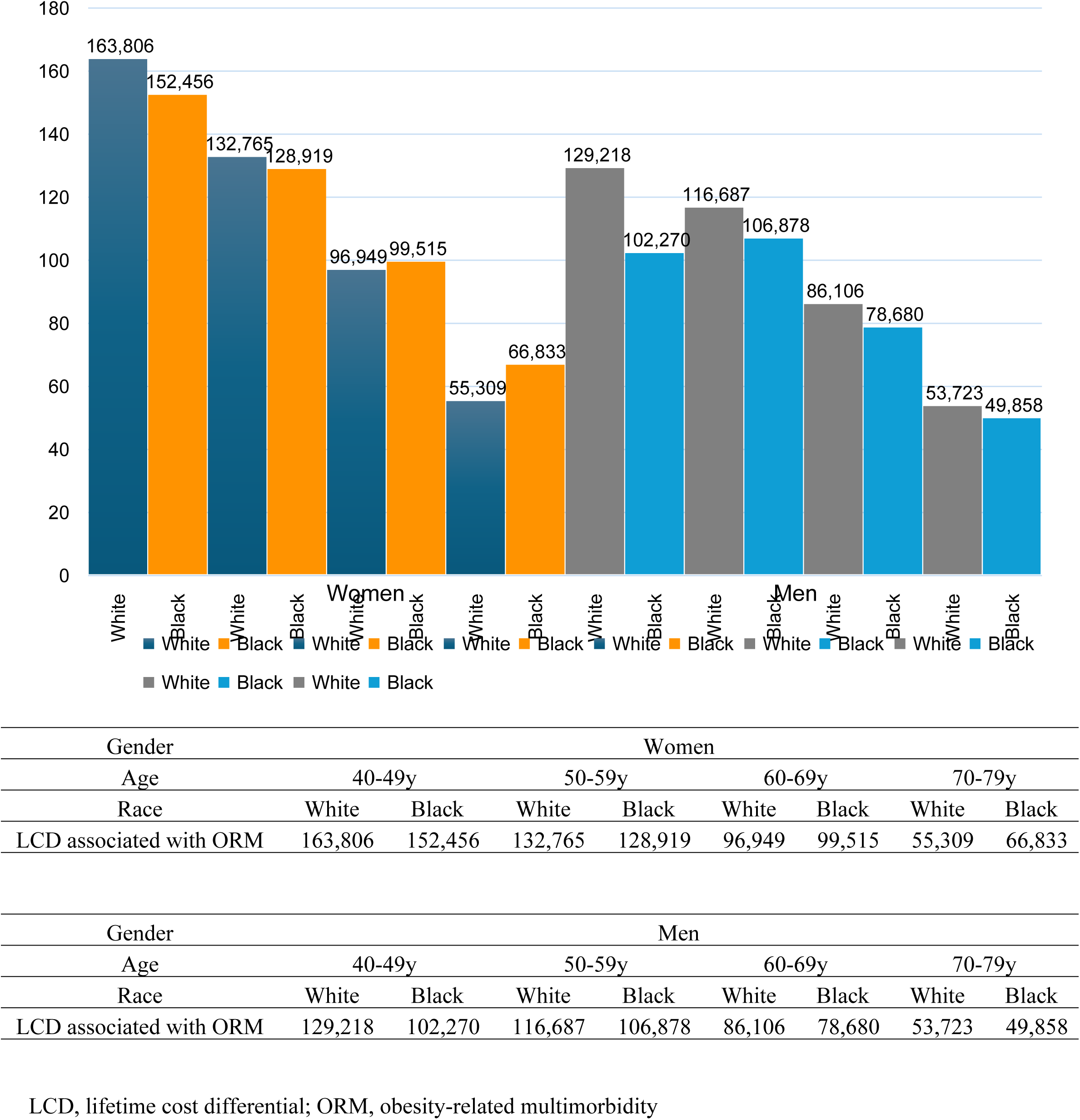
Panel A. LCD associated with ORM by age, gender, and race.

**Figure 2.**
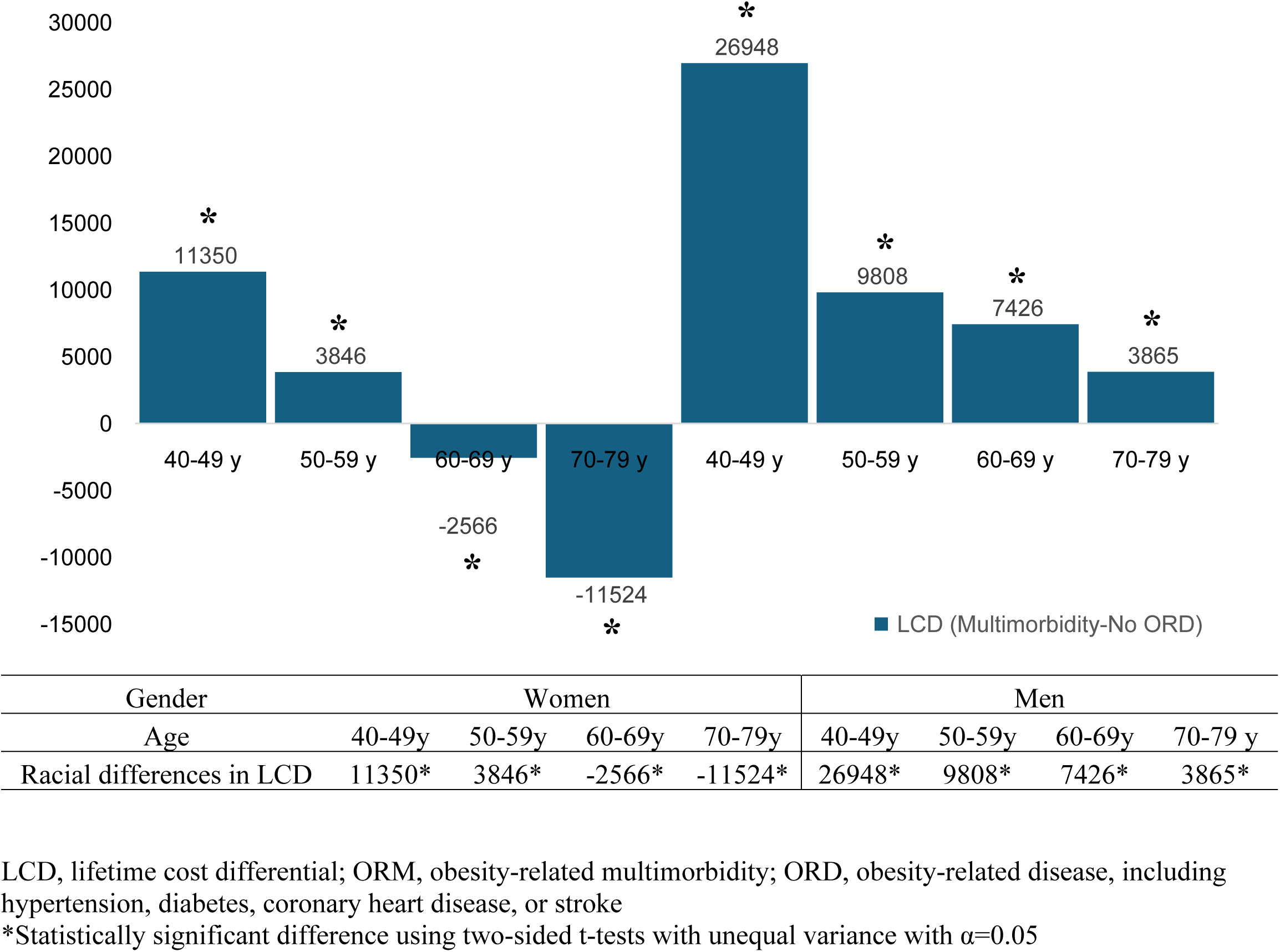
Panel B. Racial differences in LCD associated with ORM by age and gender.

## DISCUSSION

In this study we examined racial differences in LHC’s and in LCDs associated with ORM by gender and age in the U.S. general population age ≥40 years. While healthcare costs have been a widely accepted measure of population health burden,^38^ to our knowledge, this study is the first to explore lifetime healthcare costs based on ORM status and racial difference in these costs in the middle-aged U.S. general population. We found that racial differences in LHC’s were significant in all gender-age-ORD status groups, especially women and men of younger ages with ORM. We also found that racial differences in LCDs associated with ORM were prominent in all gender-age groups, particularly, LCDs for the White populations in men of all ages and women of younger ages were larger than those for their Black counterparts, while opposite in women of older ages. The study findings will deepen our understanding of the economic burden of ORM in U.S. populations of White and Black races and will inform public health policies of the subpopulations that should be prioritized to improve health disparities in obesity and ORM and the resulting financial toxicity.

While the racial differences in LHCs exist in all gender-age-ORM status groups, we found that generally the differences were more pronounced in the groups with ORM with the exception of women aged 60-69 years. Furthermore, the differences were contributed from higher LHC’s in the White populations than those in the Black populations with the exception of women aged 70-79 years. LHC’s were computed by life years and annual healthcare costs managing the existing ORDs in the year. Therefore, other things being equal, LHC’s decreased with age but increased with ORM. Previous studies also found that annual healthcare costs were higher in women than in their men counterparts and in White populations than in their Black counterparts. All of these contributed to the synthesized patterns demonstrated in the racial difference in LHCs. Our findings in all gender-age-ORD status groups are consistent with previous findings. However, our findings were able to identify the heterogeneity in racial difference in certain subpopulations, which warrants further investigation and attention.

Interestingly racial difference in LCD associated with ORM (LCD_W_-LCD_B_) decreased with age in men and women. However, we note that for women in the older age groups, LCDs associated with ORM for the White populations were smaller than those for the Black populations. There are several reasons that could explain the observed heterogeneity in racial differences in LHC’s and LCDs associated with ORM across subpopulations. First, the consequences of obesity may take years to develop and may accelerate multiple chronic health conditions and declines in overall systemic health in black women. Second, structural and socio-contextual factors, such barriers to housing, transportation, lower education and income, access to timely health care and treatment along with experiences of racism and these psychological stressors can get under the skin among, i.e., Black women imposing a higher risk for accumulation of multiple chronic conditions. This cumulative disadvantage of multimorbidity over lifespan may give rise to divergent LHC’s between Black and White older women.^8,9^ Third, oldest black women may also get delayed diagnosis and delayed follow-up care and treatment for ORD which can result in higher economic expenditures for unnecessary tests and treatments.^33^

Furthermore, our previous study on racial disparity in life expectancy in Whites vs. Black populations found younger White populations, i.e., 40-69 years to have higher life expectancy (LE) or life expectancy advantage with women to have higher LE than men, but the advantage in LE seemed to disappear for older White women, i.e., 70-79 years, which suggests gender and age contribute to differential LE, in LHC, and in LCD. Other prior research suggests Black persons have lower prevalence and risks for CHD in comparison to White persons.^23,39–41^ Moreover, Safford and colleagues found Black men had a lower risk for CHD, while Black women had a higher but nonsignificant risk for incident nonfatal CHD compared to their White counterparts, which likely translates to observed racial LHC’s difference and LCD differential for multimorbidity among White vs. Black populations and among men vs. women.^16^

Our analysis has several strengths. First, our findings are coherent across studies showing racial disparity in the presence of ORM translating to differences in LE and life years lost with Bbeing at the disadvantage of such metrics.^23^ Another prior study also suggested higher morbidity and mortality rates translating to higher costs of care in patients compared to those who do not have the disease.^42^ Second, our study provides lifetime economic burden rather than risk or cost estimates at one point in time. Third, we utilized published Markov models to infer the life expectancy and the corresponding lifetime health care expenditures over cohorts of different age-race-gender-ORM status groups, which had not been reported before. Last but not least, instead of adapting estimates from existing studies, which requires stringent assumptions, our model was populated by probabilities that were consistently estimated from the same sources of nationally representative data.

There are several limitations to our study. First, we did not aim to identify racial differences due to biology. Rather, we sought to combine both socioeconomic and biological differences in the racial differences and differential we estimated. As a result, we did not include socioeconomic variables in our cost analyses. Since socioeconomic status can fall along the causal pathway linking race and health, adjusting it therefore may not give a complete picture of the full extent of the difference.^43,44^ Second, the race category did not account for ethnicity since grouping race by ethnicity would have divided our racial groups into more than three race/ethnic groups thereby resulting in no data for respondents for several age, gender, race/ethnic BMI subpopulations with different health states. We evaluated several analytic considerations to weigh the benefit of adding ethnicity vs. the drawback of reducing the number of age groupings. For example, pooling a greater number of MEPS data or grouping age in less than four age categories. We discovered pooling additional data would not be beneficial since age of diagnosis for our target conditions was not available in MEPS until 2008. Given the prevalence of multiple chronic conditions varies across age cohorts, we decided to not reduce the number of age cohorts. Upon careful consideration, we focused our study population to two racial groups and decided to examine White vs. Black population difference and differential by age and gender. Nevertheless, we acknowledge the potential impact of not omitting the Hispanic population from our race category and identified 14.2% of the White population and 1.8% of the Black population in our analytic cohort to be Hispanic persons. We recognize this impact may not be negligible if Hispanic White population in our study significantly differed from non-Hispanic White population. Third, MEPS data, like other nationally representative data, is self-reported. Prior research has demonstrated self-reported data to be reasonably accurate, however, our estimates on healthcare costs could be biased due to non-random reporting error. Fourth, our study design was observational and causality cannot be delineated. Nevertheless, MEPS being the most complete source of data on healthcare costs and healthcare use allowed us to estimate healthcare costs for a large population of 108 million middle-aged and older U.S. adults. Fifth, due to limited availability of data on other diseases, we could only include four ORDs. We recognize this could potentially overestimate disease burdens given other costly conditions have not been accounted for in our estimation. However, we consider the bias of including four conditions to be smaller in comparison to the reported bias if we were to include only one target condition. Fifth, MEPS did not allow us to differentiate type 1 vs. type 2 diabetes. Lastly, our study findings are limited to those with a diagnosis of diabetes and may not be generalizable to the nearly 3.7 Americans with undiagnosed diabetes in 2010 or to the black population, who may have a higher burden of undiagnosed diabetes or prediabetes.^28^

## CONCLUSIONS & POLICY IMPLICATIONS

Our study provides evidence for significant racial differences in LHCs among all gender-age-ORD status subpopulations and in LCD associated with ORM among all gender-age subpopulations in the U.S. population aged 40 years or older. We specifically quantified the financial burden of ORM for each subpopulation in terms of LHC’s and LCDs associated with ORM and differences across the Black and the White populations to help inform public health policies.

Future studies investigating innovative models of integrated care utilizing community-clinical partnerships for preventing and managing multimorbidity especially in the vulnerable and at-risk Black populations may be crucial to mitigate racial disparities in financial toxicity. Such interventions could potentially benefit from culturally sensitive strategies to mitigate obesity prevalence and ORM by incorporating social context, such as neighborhood level, education, wealth, racial/ethnic composition to promote timely access to care and improved health outcomes.^43,45,46^ Future research on the social and structural determinants of health is critical to reduce the higher economic burden of obesity-related multimorbidity.^47,48^

## Data Availability

All data produced in the present work are contained in the manuscript

https://meps.ahrq.gov/mepsweb/

## ACKNOWLEDGEMENTS

This work is accepted for presentation at the 2022 National Institutes of Health’s (NIH) / National Institutes of Diabetes and Digestive and Kidney Diseases (NIDDK) Network of Minority Health Research Investigators (NMRI) Annual Meeting, April 20-22.

## Disclaimer

The conclusions and opinions presented herein are solely the responsibility of the authors and do not necessarily represent the official views of the Foundation for Barnes-Jewish Hospital, the National Institutes of Health or the Agency for Healthcare Research and Quality.

## Prior presentations

Preliminary work was presented at the 2022 International Society of Pharmacoeconomics & Outcomes Annual Scientific Meeting and the 2022 Network on Minority Research Investigators Workshop of the National Institutes of Diabetes and Digestive and Kidney Diseases.

---

**Supplementary Figure 1:**
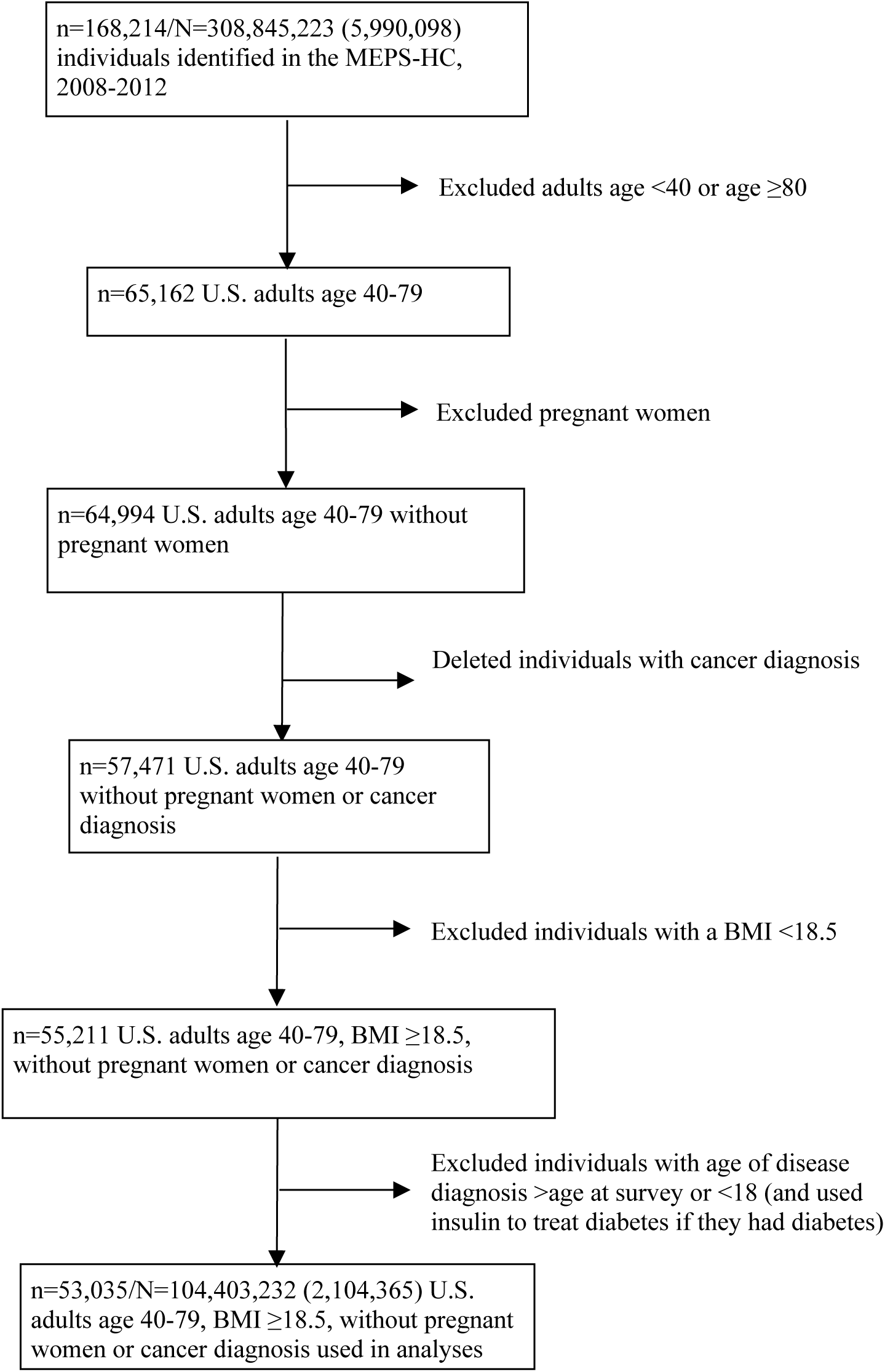
Data attrition diagram, MEPS, 2008-2012.

